# *FBXO22* deficiency defines a pleiotropic syndrome of growth restriction and multi-system anomalies associated with a unique epigenetic signature

**DOI:** 10.1101/2024.09.28.24314530

**Authors:** Navin B. Ramakrishna, Yoshikazu Johmura, Nur Ain Ali, Umar Bin Mohamad Sahari, Malak Alghamdi, Peter Bauer, Suliman Khan, Natalia Ordoñez, Mariana Ferreira, Jorge Pinto Basto, Fowzan S. Alkuraya, Eissa Ali Faqeih, Mari Mori, Naif A. M. Almontashiri, Aisha Al Shamsi, Gehad ElGhazali, Hala Abu Subieh, Mode Al Ojaimi, Ayman W. El-Hattab, Said Ahmed Said Al-Kindi, Nadia Alhashmi, Fahad Alhabshan, Abdulaziz Al Saman, Hala Tfayli, Mariam Arabi, Simone Khalifeh, Alan Taylor, Majid Alfadhel, Ruchi Jain, Shruti Sinha, Shruti Shenbagam, Revathy Ramachandran, Umut Altunoğlu, Anju Jacob, Nandu Thalange, Jay W. Shin, Almundher Al-Maawali, Azza Al-Shidhani, Amna Al-Futaisi, Fatma Rabea, Ikram Chekroun, Mohamed Al Marri, Tomohiko Ohta, Makoto Nakanishi, Alawi Alsheikh-Ali, Fahad R. Ali, Aida M. Bertoli-Avella, Bruno Reversade, Ahmad Abou Tayoun

**Author notes:** Joint senior authors to whom correspondence should be addressed: Bruno Reversade, Aida M. Bertoli-Avella, Ahmad Abou Tayoun.

## Abstract

*FBXO22* encodes an F-box protein which acts as a substrate-recognition component of the SKP1-CUL1-F-box (SCF) E3 ubiquitin ligase complex. Despite its known roles in the post-translational ubiquitination and degradation of specific substrates, including histone demethylases, the impact of FBXO22 on human development remains unknown. Here, we characterize a pleiotropic syndrome with prominent prenatal onset growth restriction and notable neurodevelopmental delay across 14 cases from 12 families. Through exome and genome sequencing, we identify three distinct homozygous loss-of-function *FBXO22* variants segregating with the disease: p.(Arg53Serfs*13), p.(Pro3Leufs*3) and p.(Val240Alafs*6), all predicted to lead to premature translation termination due to frameshift effects. We confirm that patient-derived primary fibroblasts are bereft of FBXO22 and show increased levels of the known substrate histone H3K9 demethylase KDM4B. Accordingly, we delineate a unique epigenetic signature for this disease in peripheral blood. Altogether, we identify and demonstrate that FBXO22 deficiency leads to a pleiotropic syndrome in humans encompassing growth restriction and neurodevelopmental delay, the pathogenesis of which may be explained by broad chromatin alterations.

## MAIN

Ubiquitin-tagged proteasomal degradation of specific proteins is an essential molecular process that contributes to protein turnover, thus regulating many cellular processes, including cell growth, proliferation, and differentiation.^1–3^ When the ubiquitin-proteasome system is impaired, the resultant aberrant stabilization of protein substrates can lead to defects in both common diseases such as cancer, as well as in development, leading to rare genetic diseases.^4–7^ Ubiquitin tagging of protein substrates is performed by several E3 ubiquitin ligases in complex with accessory proteins - one of which includes the SKP1-CUL1-F-box (SCF) RING-finger E3 ubiquitin ligase complex. A family of proteins termed the F-box proteins interact with the SCF via their F-box domains, functioning as the variable substrate-recognition component of the SCF. Three classes of F-box proteins exist classified according to the substrate-recognition domains present: FBXW (WD40 repeat domains), FBXL (leucine-rich repeat) and FBXO (other non-fully characterized domains).^8–10^

FBXO22 is one of at least 40 FBXO proteins^9,11^ annotated in the human genome that is ubiquitously expressed and has been characterized to play a role in the regulation of cancer.^12,13^ In particular, FBXO22 has been identified as a regulator of senescence, as well as a promoter of breast and lung cancer during early oncogenesis, while a suppressor of migration and metastasis during late cancer stages, through the interaction with its identified substrates such as KDM4A, KDM4B, TP53, PTEN and KLF4.^12–20^ In addition, its interaction with SKP1 of the SCF E3 ligase complex has been experimentally validated.^15,18,20^ While a role for FBXO22 in human development has not yet been described, loss-of-function variants of other family members including *FBXO7* (MIM605608), *FBXO11* (MIM607871) and *FBXO31* (MIM606604) have been shown to cause Mendelian diseases - Parkinson Disease 15 (PARK15, autosomal recessive, MIM260300),^21–23^ Intellectual Developmental Disorder with Dysmorphic Facies and Behavioral Abnormalities (IDDFBA, autosomal dominant, MIM618089)^24–26^ and Intellectual Developmental Disorder, autosomal recessive-45 (MRT45, MIM615979),^27^ respectively. Separately, a mouse *Fbxo22* knockout model has been characterized by gross and severe growth reduction to around half the size of wild-type littermates, alongside low-penetrant postnatal lethality.^14^

Here, we identify a pleiotropic syndrome with prominent early-onset growth restriction and notable neurodevelopmental delay across 14 cases - comprising 13 individuals and one fetus - from 12 families spanning three countries. Through a mixture of exome and short and long-read whole genome sequencing, we identify three distinct homozygous germline *FBXO22* coding variants segregating with the disease following an autosomal recessive mode of inheritance. All three alleles are expected to lead to premature translation termination and nonsense-mediated decay arising from frameshift mutations. In vitro analysis of a patient-derived primary fibroblast line confirmed that the absence of FBXO22 led to an increase in protein levels of a critical epigenetic protein substrate, while peripheral blood DNA methylation analysis identified a unique epigenetic signature. We propose that these biallelic loss-of-function (LoF) mutations of *FBXO22* lead to an aberrant stabilization of protein substrates as the cause of this previously undescribed pleiotropic syndrome.

In a collaborative international effort of clinicians and scientists, we identified 13 affected children (eight females and five males) and one fetus (14 cases in total) presenting a common core symptomatology of early onset growth restriction, neurodevelopmental delay, craniofacial abnormalities and additional poly-malformations (cardiovascular, gastrointestinal, urinal and endocrinal) (Figure 1A). All patients belonged to 12 families from three Gulf Cooperation Council (GCC) countries (), of which 10 were identified as consanguineous. Of the 14 cases, two passed away (F7-II:1 and F9-II:1) and one was a second-trimester Termination of Pregnancy (TOP) of unknown sex (Figure 1 and Table S1).

**Figure 1.**
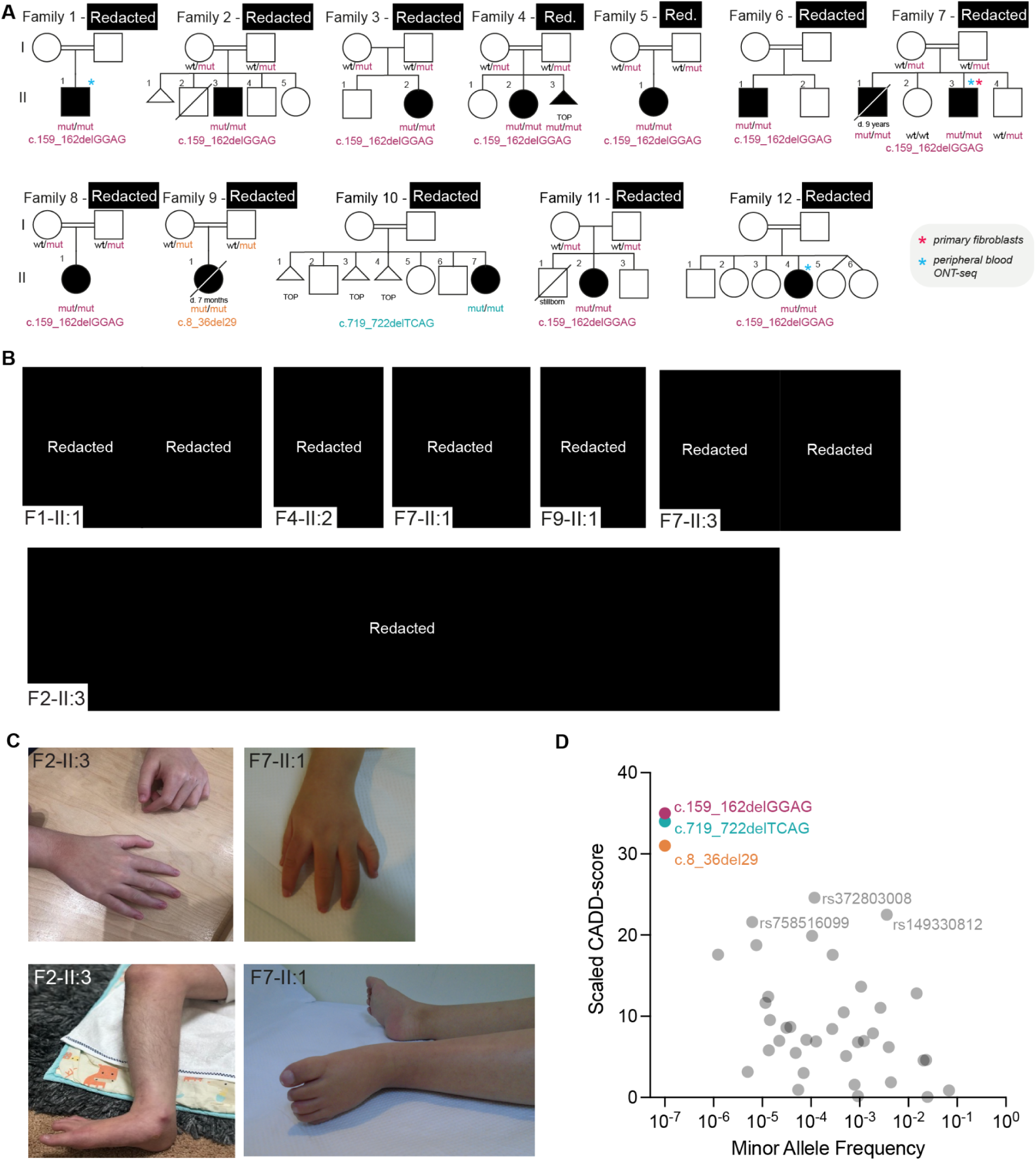
Predicted biallelic loss-of-function *FBXO22* variants in 14 cases with multi-system anomalies. (A) Pedigrees of 12 families segregating autosomal recessive congenital multi-system anomalies. Crossed symbols indicate deceased individuals. Triangular symbols indicate Termination of Pregnancy (TOP) or miscarriage. Germline *FBXO22* variant coordinates are indicated below the pedigrees, colored by variant. (B) Facial images of four affected individuals (top row, left), and timelapse facial images of individuals F7-II:3 (top row, right), and F2-II:3 from (bottom row). (C) Images of hands and feet featuring the tapering digits of affected individuals F2-II:3 and F7-II:1. (D) Minor allele frequency (x-axis) and scaled-CADD score (y-axis) of homozygous *FBXO22* coding variants found in gnomAD v.4.0.1 (grey dots, n = 37) and those found in each of the 12 families coloured by variant (n = 3). Common variants with scaled-CADD score >20 are labelled.

The 13 affected children who were clinically assessed presented with a severe growth retardation phenotype with intrauterine growth restriction (HP:0001511) (69.2%), short stature (HP:0004322) (61.5%), decreased body weight (HP:0004325) (53.8%), and an overall failure to thrive (HP:0001508) (84.6%) (Table 1 and Table S1). In addition, an apparent neurodevelopmental phenotype was observed, with the vast majority of individuals suffering from neurodevelopmental delay (HP:0012758) (92.3%), together with microcephaly (HP:0011451) (69.2%), intellectual disability (HP:0001249) (46.2%), muscular hypotonia (HP:0001252) (61.5%), generalized hypotonia (HP:0001290) (46.2%), frequent seizures (HP:0001250) (53.8%) and poor suck (HP:0002033) (38.5%) (Table 1 and Table S1). Furthermore, stark yet similar abnormal craniofacial abnormalities (HP:0001999) were observed across the individuals (84.6%), as demonstrated here in images of six individuals (obtained with parental consent) (Figure 1B). The craniofacial phenotype included a high forehead (HP:0000348) (53.8%), depressed nasal bridge (HP:0005280) (61.5%) and short nose (HP:0003196) (30.8%), hypertelorism (HP:0000316) (46.2%) with short palpebral fissure (HP:0012745) (30.8%), low-set ears (HP:0000369) (38.5%), and a narrow palate (HP:0000189) (30.8%) (Figure 1B, Table 1 and Table S1). The facial phenotype in affected individuals may show temporal evolution, most evident in a three-year-long and a 12-year-long facial photograph time-lapse for patients F7-II:3 and F2-II:3, respectively (Figure 1B). In infancy, the face tends to be round with sparse eyebrows and bears a superficial resemblance to that observed in type II collagen defects due to a depressed nasal bridge and a short nose. Over time, the face becomes triangular and eyebrows become horizontal with a downward lateral curve, while still demonstrating medial sparsity. This distinctive eyebrow morphology may provide a diagnostic handle for the *FBOX22*-related phenotype in older children.

**Table 1.** Summary of key clinical data of patients with biallelic *FBXO22* LoF variants.

| Key clinical feature of *FBXO22* deficient patients | Standardized Human Phenotype Ontology | Penetrance |
| --- | --- | --- |
| Total number of affected patients |  | 13<sup>a</sup> |
| ***Growth*** |  |  |
| Failure to thrive | HP:0001508 | 84.6% |
| Intrauterine growth restriction | HP:0001511 | 69.2% |
| Short stature | HP:0004322 | 61.5% |
| Decreased body weight | HP:0004325 | 53.8% |
| ***Neurodevelopment*** |  |  |
| Neurodevelopmental delay | HP:0012758 | 92.3% |
| Microcephaly | HP:0011451 | 69.2% |
| Muscular hypotonia | HP:0001252 | 61.5% |
| Seizures | HP:0001250 | 53.8% |
| Intellectual disability | HP:0001249 | 46.2% |
| Generalized hypotonia | HP:0001290 | 46.2% |
| Poor suck | HP:0002033 | 38.5% |
| ***Craniofacial*** |  |  |
| Abnormal craniofacial shape | HP:0001999 | 84.6% |
| Depressed nasal bridge | HP:0005280 | 61.5% |
| High forehead | HP:0000348 | 53.8% |
| Hypertelorism | HP:0000316 | 46.2% |

<sup>a</sup>Excluding fetal case (F4-II:3) where full phenotyping was not possible.

Although the growth impairment, neurodevelopmental delay and craniofacial anomalies were the most apparent, additional phenotypes with clinical variability were also observed. Cardiovascular defects were noted with atrial septal defect (HP:0001631) (30.8%) and patent ductus arteriosus (HP:0001643) (30.8%), as well as gastrointestinal defects with gastroesophageal reflux (HP:0002020) (38.5%), duodenal atresia (HP:0002247) (38.5%) and feeding difficulties (HP:0011968) (30.8%) (Table S1). Several individuals additionally showed skeletal presentations with hip dislocation (HP:0002827) (30.8%), as well as camptodactyly (HP:0100490) (23.1%) with tapered digits (HP:0001182) (23.1%), as demonstrated by individuals F2-II:3 and F7-II:1 (Figure 1C and Table S1). The fingers of these two individuals showed a resemblance to that characteristic of Coffin-Lowry syndrome (MIM303600), in exhibiting a soft appearance with distal tapering. Some of the affected individuals also showed variable endocrine abnormalities, including hyperthyroidism (HP:0000836) (7.7%) and hypothyroidism (HP:0000821) (23.1%) (Table S1).

To identify the underlying genetic cause of this pleiotropic syndrome, we performed either exome sequencing or whole genome sequencing on the affected individuals from all 12 families (Table S1). Mendelian recessive inheritance was favored across the largely consanguineous backgrounds of the families, with no sex-linked segregation observed, allowing us to focus on autosomal homozygous variants among the affected children.

Three distinct germline recessive variants in *FBXO22* (MIM609096) were identified across the 12 families, with full segregation observed in families that were tested (8/8) (Figure 1A). The majority of patients (families F1-8, F11-12) had the homozygous allele c.159_162delGGAG, while F9-II:1 bore the homozygous allele c.8_36del29 and F10-II:7 had the homozygous c.719_722delTCAG allele. All three variants have not been annotated in public databases (gnomAD v4.1.0 - Figure 1D, ExAC, BRAVO/TOPmed) in either the heterozygous or homozygous states, alluding to their rarity.

*FBXO22* spans 7 exons, encoding the 403-amino-acid long FBXO22 protein comprising three domains: an N-terminal SCF-interacting F-box domain, the protein substrate interacting central F-box and Intracellular Signal Transduction (FIST)-N domain, and C-terminal FIST-C domain (Figure 2A).^9,13,14,18,28^ All three variants encode frameshift alleles predicted to result in premature termination codons (PTCs) - c.8_36del29; p.(Pro3Leufs*13), c.159_162delGGAG; p.(Arg53Serfs*13), c.719_722delTCAG; p.(Val240Alafs*6). All variants showed the highest combined annotation-dependent depletion (CADD)^29^ scores of within 31 to 35 (Figure 1D), indicating that they are predicted to be highly deleterious. In particular, all three frameshift variants are anticipated to lead to nonsense-mediated decay (NMD) due to the presence of PTCs in exon 1, 2 and 6, respectively, of the 7-exon transcript, with the new PTC of c.719_722delTCAG; p.(Val240Alafs*6) located 77 bp upstream from the final exon-exon junction, also meeting the criteria for NMD (having a PTC within the upstream exons, and >55 bp away from the final exon-exon junction if within the penultimate exon).^30–32^ In the theoretical absence of NMD, deleterious truncating mutants are expected to result with both p.(Pro3Leufs*13) and p.(Arg53Serfs*13) occurring at the N-terminus, and p.(Val240Alafs*6) truncating part of the substrate-recognizing FIST-N domain while removing the whole FIST-C domain (Figure 2A). In addition, both Arg53 and Val240 are highly conserved residues (Figure S1A).

**Figure 2.**
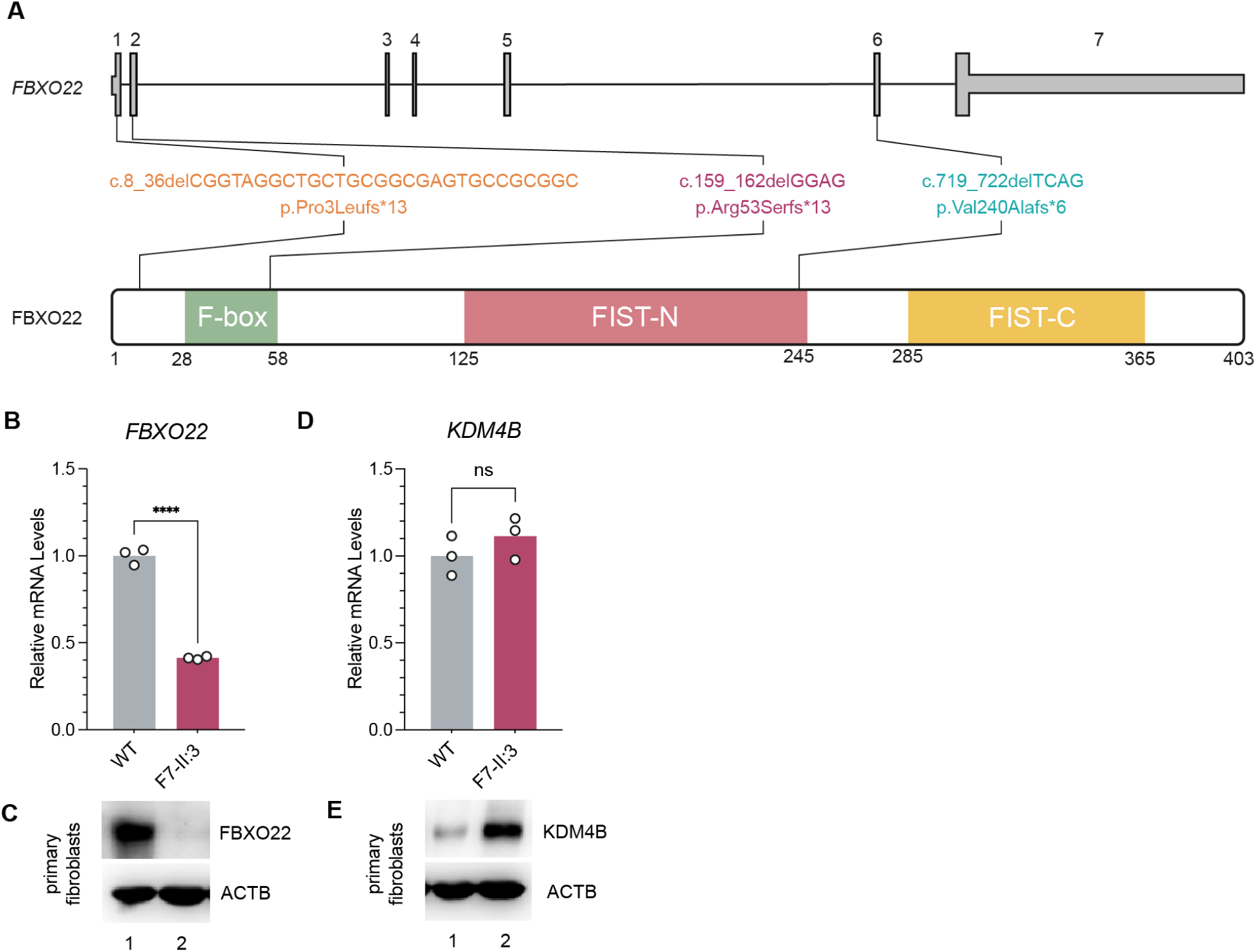
Loss of FBXO22 in patient-derived primary fibroblasts leads to abnormally high KDM4B levels. (A) Schematic diagram of the genomic (top) and protein (bottom) structure of FBXO22 in humans. The FBXO22 protein contains three conserved domains: F-Box, FIST-N and FIST-C. The three homozygous genetic variants and corresponding protein mutations are indicated. (B-E) FBXO22 and KDM4B expression analysis at the mRNA and protein levels in primary dermal cutaneous fibroblasts from F7-II:3 with control (WT). (B,D) *FBXO22* and *KDM4B* mRNA levels normalised to housekeeping gene *ACTB* mRNA levels, relative to the WT control (n = 3 biological replicates), with significant reduction in *FBXO22* levels and unaltered *KDM4B* levels. ****p = 0.0000295; ns, nonsignificant (unpaired, two-way student’s t-test). (C,E) Western blot of endogenous FBXO22 and KDM4B proteins with ACTB (beta-Actin) as the housekeeping control, showing negligible FBXO22 levels and increased KDM4B levels in the patient fibroblasts.

Three additional previously annotated homozygous alleles within *FBXO22* were observed at medium frequencies (>10^-5^) in gnomAD v4.1.0 with moderate CADD scores between 20 and 25 (Figure 1D). The single nucleotide polymorphisms (SNPs) rs758516099 (c.286C>G; p.(Arg96Gly)), rs149330812 (c.302C>T; p.(Thr101Ile)) and rs372803008 (c.631C>G; p.(Leu211Val)) all encode missense variants. In particular, a positively charged amino acid was substituted for a neutral amino acid in p.(Arg96Gly) in a loop domain between the F-box and FIST-N domains, a mildly-polar amino acid substituted for a hydrophobic amino acid in p.(Thr101Ile) at the beginning of the FIST-N domain, and a relatively equivalent hydrophobic swap of amino acids in p.(Leu211Val) within the FIST-N domain (Figure S1A). All three substitutions appear to be able to be accommodated within highly-confident Alphafold3^33^ predicted structural regions of FBXO22 - Arg96 in protein surface polar interactions (no salt bridges identified), Thr101 in tertiary β-β fold interactions and Leu211 in tertiary hydrophobic interactions in an ɑ-β loop region (Figure S1B). Altogether, as all three of these pre-annotated variants are present in population databases at frequencies more than expected for disease with no known clinical association, and with their potential accommodation in protein structure, they are unlikely to be deleterious.

To assay the pathogenicity of the frameshift variants in the ubiquitously expressed *FBXO22* (GTEx, Figure S2A), we derived primary cutaneous fibroblasts from patient F7-II:3 bearing homozygous alleles of the most common variant in our cohort (c.159_162delGGAG; p.(Arg53Serfs*13)), which was additionally verified by Sanger sequencing. As the variant was predicted to result in a PTC in an early exon, which probably leads to NMD, an RT-qPCR analysis was performed on cDNA extracted from the affected fibroblasts alongside previously derived unaffected wildtype (WT) primary fibroblasts.^34^ This analysis demonstrated a significant 2.5-fold reduction in *FBXO22* mRNA levels, down to 40% of the amount seen in the control WT fibroblasts (Figure 2B). In addition, Western blotting analysis demonstrated that endogenous FBXO22 protein levels were completely absent in the cellular extracts of the patient fibroblasts compared to the WT control (Figure 2C). These results indicate that this frameshift mutant variant likely destabilizes *FBXO22* mRNA via NMD and behaves as a LoF protein-null allele, in agreement with the above predictions.

As FBXO22 is a characterized substrate-recognition partner of the E3 ubiquitin-ligase SCF complex with known protein targets subject to ubiquitin-tagged proteasomal degradation, we investigated the impact of the loss of FBXO22 on its protein substrates. In particular, we investigated the key known substrate - the ubiquitously expressed epigenetic histone H3K9me3/2 demethylase KDM4B^13,19,35,36^ (GTEx, Figure S2B) via RT-qPCR and Western blotting of fibroblast extracts. While the mRNA expression levels of *KDM4B* in the patient cells were unperturbed compared to the WT control (Figure 2D), a stark increase in KDM4B protein expression levels was observed in the patient-derived fibroblast line (Figure 2E). This result highly suggests that KDM4B protein levels were post-translationally stabilized in the absence of SCF^FBXO22^ proteasomal-degradation activity, with no change to the upstream transcription of its gene.

Given the observed altered protein levels of the histone demethylase KDM4B, which suggest changes to chromatin in the absence of FBXO22, we turned our attention to profiling epigenetic changes in our patient samples. Notably, loss of function variants in *KDM4B*, associated with the intellectual development disorder MRD65 (MIM609765), have been previously associated with a robust DNA methylation epigenetic signature in peripheral blood.^37–39^ Additional unique DNA methylation epi-signatures have also been identified in Mendelian disorders caused by mutations in other histone demethylases and methyltransferases, with general changes in histone modifications also previously shown to impact DNA methylation.^40–43^ We therefore investigated the changes to the DNA methylomes of peripheral blood gDNA from three affected patient samples from families F1, F7 and F12 with the c.159_162delGGAG; p.(Arg53Serfs*13) variant using long read Oxford Nanopore (ONT) sequencing with 5-methylcytosine basecalling.

Focusing our DNA methylation analysis across 3,643 genomic loci corresponding to probe regions of previously identified epi-signatures encompassing 34 Mendelian neurodevelopmental disorders (Episign MNDDs),^39,44,45^ we observed that all three *FBXO22* samples formed a distinct cluster, segregating away from the other 34 Episign MNDDs as well as the control, implicating a unique *FBXO22* epi-signature (Figure S3A). We iteratively identified the top 40 differentially-methylated loci within these regions representing a proposed *FBXO22-*specific epi-signature (see Methods and Table S2).^45^ This primarily consists of marked hypomethylation (Figure 3A and Table S3), with the *FBXO22*-deficiency samples forming a highly specific cluster relative to the 34 Episign MNDDs and control in principal component analysis of methylation values of these loci (Figure 3B). Analysis of the 40 regions revealed differential methylation within specific genes or proximal regulatory elements upstream of genes (20 regions out of 40) (Table S2). Altogether, these genes are found to be predominantly active in the brain (12/20 genes), respiratory system (6/20 genes), gastrointestinal tract (5/20 genes), muscle tissues (8/20 genes), and bone marrow (4/20 genes), and have been associated with neurological and developmental disorders (12/20 genes),^46–50^ cardiovascular and blood disorders (6/20 genes)^51,52^ and various forms of skeletal abnormalities (4/20 genes)^47,53^ (Table 2). A patent finding is significant hypomethylation within the ultimate exon 19 of *AGAP2* coding for its 3′-UTR in comparison to both the EpiSign MNDD cohort as well as an additional sample set of unrelated neurological disorders (ND cohort), previously generated using the same ONT-seq long read sequencing protocol (n=17)^45^ (Figure 3C,D). Hypomethylation at the *AGAP2* 3′-UTR has previously been associated with haploinsufficiency of the H3K4 methyltransferase *KMT2D* in patients with Kabuki syndrome (MIM147920 and MIM300867)^54,55^. Notably, *FBXO22*-deficient patients and Kabuki cases present with overlapping clinical features such as developmental delay, growth failure, hypotonia and seizures.

**Figure 3.**
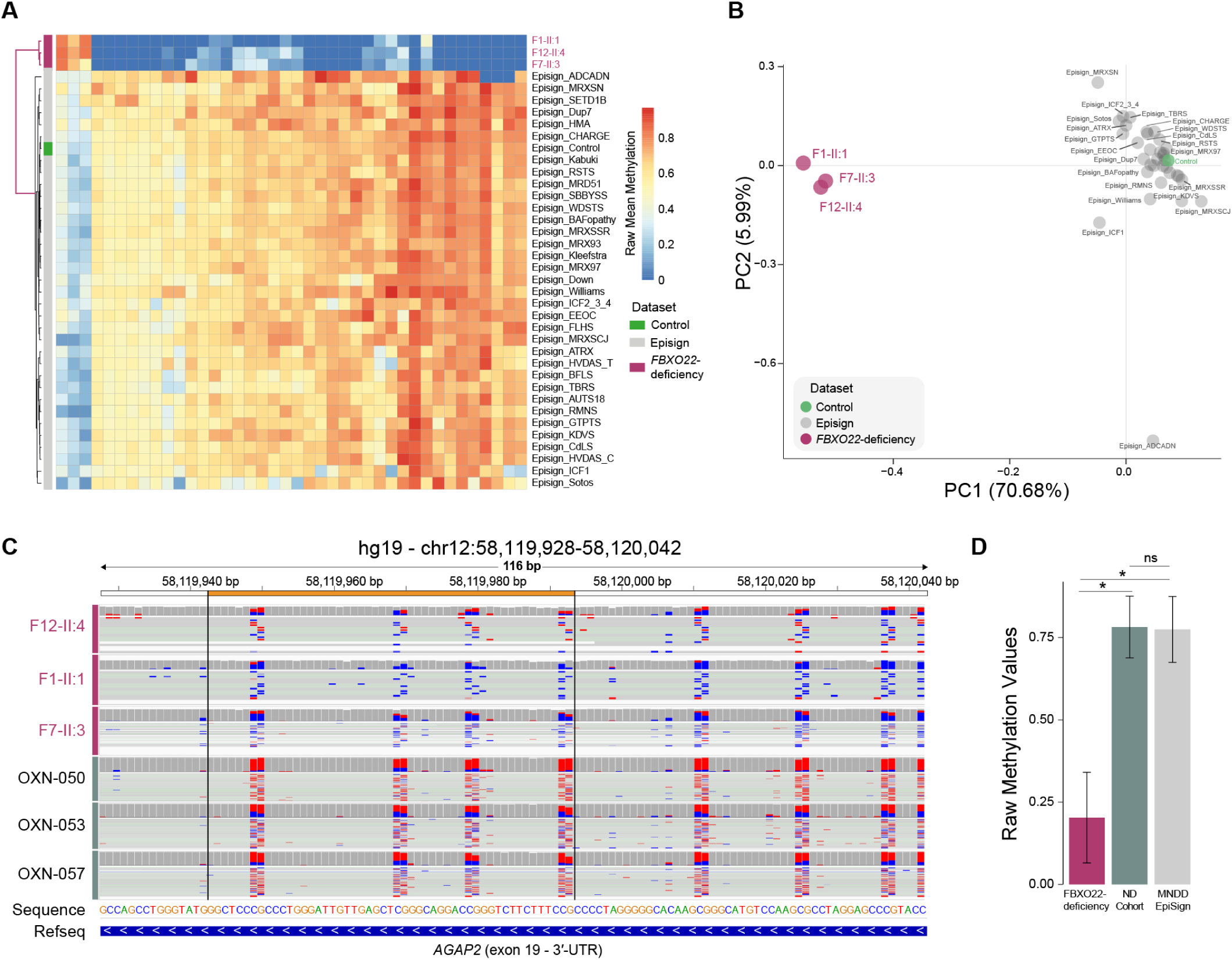
Loss of *FBXO22* is associated with a unique epigenetic signature in peripheral blood. (A) Heatmap with euclidean distance hierarchical clustering of DNA methylation values for the top 40 differentially-methylated regions featuring the *FBXO22* epi-signature for three *FBXO22*-deficiency peripheral blood samples integrated with the 34 EpiSign Mendelian neurodevelopmental disorders (MNDDs) and control dataset. (B) Principal component analysis on the 40 regions representing the DNA methylation epi-signature in PBMCs of three patients lacking *FBXO22* showing clustering of all three cases away from the EpiSign MNDDs and control. Variance explained by components PC1 and PC2 are indicated in brackets. (C) Genome browser view of the differentially methylated (orange) region of the 3′-UTR within the ultimate exon 19 of *AGAP2* featuring aggregated CpG methylation (5mC - red; unmodified C - blue) of ONT-seq long reads from the three *FBXO22*-deficiency samples and three samples from the general neurological disorders (ND) cohort. (D) Methylation values at the differentially methylated probed region within the 3′-UTR of *AGAP2* in the *FBXO22*-deficiency ONT-seq samples (n=3), ND cohort ONT-seq samples (n=17, p=0.013) and EpiSign MNDD samples (n=34, p=0.015). Error bars denote SD. *p < 0.05; ns, non-significant (unpaired, two-way student’s t-test).

**Table 2.** Expression profiles and associated diseases of genes with promoter or proximal DNA methylation differences within the top 20 differentially-methylated regions of *FBXO22*-deficiency.

| **Gene** | **Expression** | **Associated diseases** |
| --- | --- | --- |
| ***NAT8L*** | Brain – Lungs – Retina – Adipose tissue<br>– Pituitary gland – Skeletal muscle | N-acetylaspartate deficiency<br>Cardiomyopathy<br>Neurodegeneration<br>Alzheimer |
| ***DDX39B*** | Brain – Thyroid – Bone marrow –<br>Kidneys – Lungs – Skin – Heart –<br>Skeletal muscle | Neurodevelopmental delay<br>Epilepsy<br>Short stature and congenital hypotonia<br>Anterior horn cell |
| ***TRIM15*** | GI tract – Liver – Kidneys – Colon | Hepatic veno-occlusive disease with<br>immunodeficiency |
| ***TRMT9B*** | Brain – Thyroid | Dubowitz syndrome |
| ***ARHGEF10*** | Brain – Lungs – Heart – Skeletal muscle | Slowed Nerve Conduction Velocity (SNCV)<br>Axonal neuropathy |
| ***AGAP2*** | Brain | Kabuki Syndrome |
| ***GALNT9*** | Brain – Thyroid – Kidney | Association with autism spectrum disorders<br>and Parkinson's disease |
| ***ZNF385A*** | Brain – Skin – Retina | Encephalopathy<br>Cone-Rod Dystrophy 2 |
| ***OLIG1*** | Brain | Oligodendroglioma |
| ***MED13*** | Bone marrow – Liver – Lungs – Retina | MRFACD syndrome |
| ***SLC22A23*** | Brain - Parathyroid gland – GI tract | Wolf-Hirschhorn Syndrome<br>Inflammatory Bowel Disease |
| ***SLC17A5*** | Parathyroid gland – Retina – Liver | Salla disease<br>Free Sialic Acid Storage Disorders |
| ***PRDM16*** | GI tract – Thyroid – Brain – Kidney –<br>Heart | Cardiomyopathy |
| ***DDAH2*** | Heart – Lungs – Thyroid - GI tract –<br>Kidneys | Coronary artery disease |
| ***MYH7B*** | Heart – Skeletal muscles | Cardiomyopathy |
| ***MEOX1*** | Heart – Adipose tissue | Klippel-Feil syndrome |
| ***DDR1*** | Brian – Skin – GI tract – Kidneys | Spondylo-meta-epiphyseal dysplasia |
| ***LY6G6D*** | Skin – GI tract | Anemia, Nonspherocytic Hemolytic |
| ***MPIG6B*** | Lungs – Skin – Spleen | Congenital macrothrombocytopenia with focal<br>myelofibrosis |
| ***TAPBP*** | Liver – Bone marrow – Brain – Heart | Bare lymphocyte syndrome |

Separately, analysis of the methylation profile across regions previously shown to define a specific *KDM4B*-deficiency epi-signature^39^ did not show any difference for samples with biallelic *FBXO22* loss of function (n=3) in comparison with the ND cohort (Figure S3B). This result may suggest that although the known *KDM4B* epi-signature has been associated with the loss of function of *KDM4B*, the underlying DNA methylation marker loci might not be dosage-dependent to capture the opposite pattern of resultant KDM4B overexpression associated with *FBXO22* biallelic loss.

In conclusion, we have identified and characterized recessive LoF variants in *FBXO22*, which we propose are responsible for this heretofore undescribed pleiotropic congenital Mendelian syndrome. This is supported by genetic and clinical data across a large cohort of 12 families identifying three LoF alleles, target characterization using patient-derived fibroblasts in vitro, and epigenetic profiling with epimarker identification in peripheral blood gDNA.

In further support of the pathogenicity of the LoF variants, we note that *FBXO22* is intolerant to heterozygous loss-of-function variants with a very low observed/expected LoF ratio (0.402) in the general population (gnomAD v4.1.0) (Fig S4A). More importantly, given the autosomal recessive inheritance, *FBXO22* is entirely devoid of biallelic occurrences of rare (≤0.5%) LoF and/or deleterious (predicted) missense variants in gnomAD (variant co-occurrence statistics only available in v2.1.1) (Figure S4B). Interestingly, a strong founder mutation likely exists in the GCC countries from which the affected individuals originate. The c.159_162delGGAG; p.(Arg53Serfs*13) variant has a minor allele frequency of 0.05% in Qatar, where 15 heterozygous individuals were ascertained within a healthy cohort of 14,000 whole genomes. In addition, haplotype analysis of the c.159_162delGGAG; p.(Arg53Serfs*13) variant across all three ONT-seq WGS samples show that the majority of ancestry on chr15 for all haplotypes is inferred to be Middle Eastern, with minor Central & South Asian, African and East Asian ancestry (Fig S5A and B), while phylogeny analysis show that all six haplotypes (two per individual) cluster together within a larger phylogeny of haplotypes of 135 published Middle Eastern individuals (Fig S5C).^56^

Additionally, the DECIPHER database (GRCh38) reports 9 individuals with an additional copy of *FBXO22* due to duplications ranging from 0.4 to 79.4 Mb on chromosome 15q24.2. No apparent phenotypes are attributed to these cases, suggesting that three copies of *FBXO22* may not be pathogenic. Additional searches in public databases have identified *FBXO22* had no hits in public PheWAS, but some genome-wide significant hits in GWAS for kidney function (NHGRI-EBI GWAS Catalog: GRCh38.p14 and dbSNP Build 156). *FBXO22* was deemed a hit 27 times in 1,356 CRISPR screens (BioGRID ORCS v1.1.16.1), in which its targeted inhibition was often associated with decreased cellular proliferation (52% of hits). This can be interpreted to be in concordance with the gross growth reduction seen across patients and in the mouse KO model.^14^

The ubiquitous expression of *FBXO22* across tissues lends support to the pleiotropic nature of the syndrome, with the demonstrated loss of FBXO22 protein and accompanying NMD in patient-derived fibroblasts leading to an ectopic stabilization of a prototypical protein substrate. In addition to KDM4B, additional epigenetic substrates subject to polyubiquitination-targeted protein degradation by SCF^FBXO22^ have been identified to date, which include the histone demethylases KDM4A and KDM5A, histone methyltransferase NSD2, and the histone acetyl reader BRD4.^16,18,57,58^ Further characterized substrates include the tumor suppressors and cell cycle regulators KDM4A-methylated TP53 (p53), PTEN, CDKN1A (p21), CDKN1C (p57Kip2), LKB1, as well as additional targets KLF4, BACH1, HDM2, CD274 (PD-L1), CD147 and FKBP12.^14,15,17,20,58–65^ Beyond ubiquitin-mediated protein degradation, MTOR (mTOR) has additionally been identified as a non-proteolytic monoubiquitinated substrate of FBXO22, serving a regulatory role in amino acid level sensing.^66,67^ With the vast majority of studies performed in cancer cell lines, the repertoire of substrates largely discovered has thus implicated FBXO22 as an epigenetic multiplayer in carcinogenesis and therapy response, particularly as a regulator of senescence, as well as a promoter of breast and lung cancer proliferation in early cancer stages, while a suppressor of migration and metastasis during late cancer stages.^12,13,16,59^ To date, no predisposition to or protection against cancer in the probands have been documented, nor in the aforementioned mouse KO line, which instead demonstrated severe growth reduction and occasional early postnatal lethality.^14^

The observed stabilization and increase in KDM4B protein levels in patient-derived fibroblasts, together with knowledge of additional epigenetic protein targets, similarly posits FBXO22 as a potential epigenetic multiplayer in human development. As mentioned above, haploinsufficiency of *KDM4B* is causal for an autosomal dominant intellectual developmental disorder (MRD65, MIM609765), characterized by overall delayed neurodevelopment, dysmorphic facial features, feeding difficulties, and hypotonia.^68^ The overlap in neurological and additional defects in MRD65 and FBXO22-deficiency could suggest the importance of dose control of protein levels of KDM4B in neurodevelopment. In addition, biallelic pathogenic variants in the homolog *KDM5B* are also responsible for an autosomal recessive intellectual disorder (MRT65, MIM618109), with neurodevelopment similarly impacted.^69–71^ This precedence, together with the wide range of likely impacted additional protein substrates of FBXO22 across multiple tissue types, could potentially account for the neurological and the additional multi-system anomalies seen in patients, warranting further exploration in organ-specific assays.

Both recessive and dominant causative variants in lysine demethylases and methyltransferases implicated in Mendelian neurological disorders have previously been associated with peripheral blood DNA methylation changes. In addition to *KDM4B* and *KDM5B* described above, epigenetic signatures have been identified for neurological disorders implicating loss of function for *KDM2B* (*KDM2B*-related syndrome),^72^ X-linked recessive *KDM5C* (MRXSCJ, MIM300534), X-linked dominant *KDM6A* or autosomal-dominant *KMT2D* (Kabuki syndrome 1 and 2, MIM147920 and MIM300867), autosomal dominant *KMT2A* (WDSTS, MIM605130), autosomal dominant *KMT2B* (DYT28 and MRD68, MIM617284 and MIM619934), autosomal dominant *KMT2C* (Kleefstra syndrome 2, MIM617768), autosomal dominant *EHMT1* (Kleefstra syndrome 1, MIM610253) and autosomal dominant *KMT5B* (MRD51, MIM617788).^37–39^ In addition, autosomal dominant mutations in the ubiquitin ligase *FBXO11* (IDDFBA, MIM618089)^24–26^ are also marked by a DNA methylation signature (Episign v5). With FBXO22 molecularly implicated in regulating protein levels of several lysine demethylases and methyltransferases, likely impacting chromatin states, and with its association with neurological dysfunction, an epigenetic DNA methylation signature for *FBXO22*-deficiency, likely converging on a common methylation target (*AGAP2* gene) with *KMT2D*, was readily and likewise identified in peripheral blood gDNA. The *FBXO22* epi-signature presents an opportunity as a biomarker for detecting *FBXO22*-deficiency while warranting further investigation into the specific molecular epigenetic pathways perturbed.

Taken together, our clinical, genetic and molecular studies define a heretofore undescribed Mendelian recessive disorder caused by homozygous LoF *FBXO22* variants, characterized by multi-system anomalies and a unique epigenetic signature. The ubiquitous expression of *FBXO22* coupled with the extensive repertoire of protein targets, including epigenetic modulators, thus provides a potential underlying rationale for the pleiotropic effects of *FBXO22*-deficiency.

## DATA AND CODE AVAILABILITY

The data and code parameters that support the findings of this study are detailed within the methods and supplemental information, or from the corresponding authors upon request.

## SUPPLEMENTAL INFORMATION

This manuscript contains five supplementary figures, the unabridged version of Table 1 (Table S1), a supplementary table of *FBXO22*-deficiency methylation coordinates and values (Table S2), a supplementary table of annotations of the methylation coordinates (Table S3) and supplemental case reports (fully redacted).

## ACKNOWLEDGMENTS

We thank all the families for partaking in this study and the referring clinicians for their generous help. We thank the Genome Institute of Singapore Sequencing and Genotyping Platform for ONT-seq services in Singapore. N.B.R. is a recipient of Singapore Ministry of Health’s RIE2025 National Medical Research Council OF-YIRG award (OFYIRG23jan-0036; MOH-001341) administered by the Agency for Science, Technology and Research. A.A.M is a recipient of Sultan Qaboos University Strategic research funding (SR/MED/GENT/16/01). B.R. is a fellow of the Branco Weiss Foundation (Switzerland) and an EMBO Young Investigator (Europe) and is funded by BESE at KAUST in the KSA. We would also like to acknowledge the support of the Mohammed Bin Rashid University of Medicine and Health Sciences and Al Jalila Foundation.

## AUTHOR CONTRIBUTIONS

A.A.T. and B.R. designed, conceived and supervised the study.

A.M.B.-A, ascertainment, phenotype and genotype analyses of families 1 to 10.

S.K., N.O. and M.F. performed exome/genome sequencing data analyses and variant interpretation of families 1 to 10 with supervision of J.P.B., P.B. and A.M.B.-A.. A.A.-F., A.A.-Shidhani, A.A.-M, M.Alghamdi, F.S.A., E.A.F., M.M., N.A.M.A., A.A.Shamsi, G.E., H.A.S., M.A.O., A.W.E.-H., S.A.S.A.-K., N.A., F.A., A.A.Saman, H.T., M.Arabi, S.K., A.T., M.Alfadhel, R.J., S.Shenbagam, A.J., N.T., F.R., I.C., M.A.M. and A.A.-A. were involved in conducting the clinical and genetic evaluation of the patients, organisation of clinical information, the collection of human biological samples and the whole genome/exome sequencing for the affected families.

U.A. reviewed facial findings to evaluate dysmorphic features and contributed to the writing of the relevant part of the article.

N.B.R., Y.J., N.A.A., U.B.M.S., A.A.T., S.Sinha and R.R. performed experimental work, formal data analysis and visualisation with supervision from F.R.A., T.O., M.N., A.A.T. and B.R..

N.B.R., A.A.T. and B.R. wrote and edited the manuscript.

N.B.R., J.W.S., F.R.A., A.M.B.-A, A.A.T. and B.R. provided resources and acquired funding.

## WEB RESOURCES

The following web resources were used in this study: The Online Mendelian Inheritance in Man (OMIM): http://www.omim.org; The Exome Variant Server (ftp.ncbi.nlm.nih.gov/pub/clinvar/vcf_GRCh37) from NHLBI Exome Sequencing Project (ESP): http://evs.gs.washington.edu/EVS/; 1000 Genome Project Database: http://browser.1000genomes.org/index.html; Genome Aggregation Database (GnomAD v4.1.0 (hg38) and v2.1.1 (hg19)): http://gnomad.broadinstitute.org/; BRAVO/TOPmed database: (https://bravo.sph.umich.edu/freeze8/hg38/); Greater Middle East (GME) Variome web: http://igm.ucsd.edu/gme/index.php; NCBI dbSNP: http://www.ncbi.nlm.nih.gov/SNP/; DECIPHER database (GRCh38, v11.27): https://www.deciphergenomics.org; PheWAS catalog: https://phewascatalog.org/phewas; NHGRI-EBI GWAS catalog (GRCh38.p14 amd dbSNP build 156): https://www.ebi.ac.uk/gwas/; BioGRID ORCS v1.1.16.1: https://orcs.thebiogrid.org

## MATERIALS AND METHODS

### Ethical approval

Written informed consent was obtained from all individuals (parents and parents on behalf of patients from each family) for genetic testing, skin biopsy (for patient F7-II:3) and the use of the clinical information and images in this study, according to the ethical approval of the local Institutional Review Boards (IRBs) in and. The study protocol was approved by A*STAR IRB (2019-087) in Singapore and at KAUST (23IBEC090) in the KSA.

### Patient recruitment

The affected patient F1-II:1 and F12-II:4 were diagnosed by A.A.T. at. The affected patient F2-II:3 was diagnosed by A.M.A.M.A.S. in. The affected patient F3-II:2 was diagnosed by G.E. in. The affected patient F4-II:2 and fetal case F4-II:3 were diagnosed by H.A., M.A.O. and A.W.E. in. The affected patient F5-II:1 was diagnosed by S.A.S.A. at. The affected patients F6-II:1 and F8-II:1 were diagnosed by N.A. at. The affected patients F7-II:1 and F7-II:3 were diagnosed by A.A. at. The affected patient F9-II:1 was diagnosed by M.Alfadhel, F.A. and M.Alghamdi in. The affected patient F10-II:7 was diagnosed by A.A.S. and E.A.F. at. The affected patient F11-II:2 was diagnosed by M.M. in.

### Next-Generation Sequencing and Analysis

Whole genome sequencing and whole exome sequencing were performed at different research institutes according to local standard procedures.

### Whole Exome Sequencing (WES)

DNA was barcoded and enriched for the coding exons of targeted genes using hybrid capture technology (Agilent SureSelect Human All-exons-V6), as previously described.^73,74^ Prepared DNA libraries were then sequenced using Next-Generation Sequencing (NGS) technology [NovaSeq 6000 (Illumina), 150 bp paired-end, at 200X coverage]. The reads were mapped against UCSC GRCh37/hg19 by Burrows-Wheeler Aligner (BWA 0.7.12).

### Illumina-WGS

Whole genome sequencing (WGS) was done as previously described for F2, F3 and F9.^74^ Briefly, using gDNA extracted from whole blood, sequencing libraries were constructed on site using the TruSeqDNA PCR-Free Library Prep kit (Illumina) according to the manufacturer’s instructions. Paired-end sequencing was performed on the NovaSeq 6000 platform with the S1 flowcell (Illumina). The reads were mapped against UCSC GRCh37/hg19 by Burrows-Wheeler Aligner (BWA 0.7.12).

### Variant Analysis

Genome Analysis Toolkit (GATK 3.4) was used for variant calling. Variant filtration, as previously described,^74,75^ was applied to keep novel or rare variants (≤ 1%). Publicly available variant databases and an in-house database of 1562 exomes (for the population cases) were used to filter out common or benign variants. Only coding or splicing variants were considered. The phenotype and mode of inheritance (autosomal recessive) were considered. Variants of high impact or highly damaging missense, a CADD^29^ score ≥ 20 and shared between the affected individuals were prioritized. Other OMIM genes that are known to be associated with a similar phenotype were analyzed from the exome data and no pathogenic variants were identified.

### Long read sequencing, methylation calling and mapping

Long read sequencing, processing and methylation calling was done as previously described.^45^ Briefly, genomic DNA was extracted from peripheral whole blood using the QIAsymphony DSP DNA Kit (Qiagen) and QIAsymphony automated nucleic acid extraction instrument, according to the manufacturer’s instructions. For all samples, 1,000 to 4,500 ng gDNA was sheared with G-Tubes (Covaris LLC, USA) following the standard 20 kb protocol. The resulting DNA fragments were utilized for library preparation using the Ligation Sequencing Kit V14 (Oxford Nanopore, UK), according to the manufacturer’s instructions, and was sequenced on the PromethION P48 device with R10.4.1 flow cell (Oxford Nanopore, UK) as follows: 72 hours with a second library loaded at 24 hours post flow cell nuclease flush for FBXO22_F12:II_4 (N50: 11.39kb; Bases: 94.35 Gb; Approximate coverage: 31x); for the low DNA input sample, FBXO22_F1:II_1, DNA shearing was not performed and the library was sequenced for only 72 hours (N50: 14.9 kb; Bases: 34.5 Gb; Approximate coverage: 11.5x); for FBXO22_F7:II_3 the library was sequenced for 97 hours with second and third libraries loaded post-nuclease flushes at 28 hours and 52 hour timepoints (N50: 11.5 kb; Bases: 110.1 Gb; Approximate coverage: 36.7x). Base calling (with 5mC) was done using “high-accuracy base calling” (HAC) mode during the run using MinKnow distribution (v22.05.7 or v24.02.19) and Guppy/Dorado (v6.1.5 or v7.3.11). The methylation SAM tags (MM,ML) were preserved using samtools (version 1.13) for all BAM passed files and were then aligned to the human reference genome (GRCh37/hg19) using minimap2 (v2.22-r1101) using the appropriate parameters ‘`minimap2 -x map-ont -a -y`’.

### Methylation profile analysis

Methylation analysis was performed by comparing the methylation profile of the patients with those reported in literature for the Episign epigenomic signature^44^ for a total of 34 Mendelian neurodevelopment disorders (MNDDs). For the purpose of comparison, disease specific probes from Illumina Infinium methylation 450k and EPIC bead chip arrays identified as epi-signatures (EpiSign) were mapped on the human genome hg19 using pblat15 with the parameter “`-fastMap`”. In order to remove ambiguity coming from multimapping probes, those with block count of 1 with alignment length matching the probe length were selected and assessed for the downstream analysis. Aggregated methylation modification counts for each base from long read sequencing in the probe region were calculated using modbam2bed from the ‘methyl’ module of Epi2Me workflow wf-human-variation (v1.2.0). Methylation values for all samples and MNDD dataset were standardized (i) and normalized (ii) using min-max normalization using the equation below where s is the sample, p is the probe, x_p_ is the methylation value for each probe, *x*– is the mean and σ is the standard deviation, stdMethyl is the standardized methylation value and normMethyl is normalized methylation value:

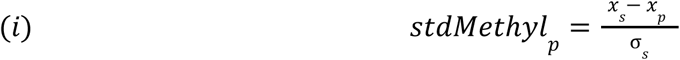

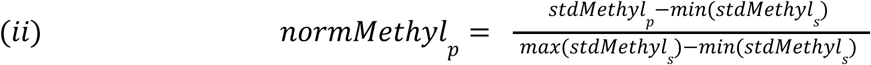

Hierarchical clustering was performed using euclidean distance and ward.D2 for MNDD epi-signature probes on normalized methylation values for each sample with the MNDD. For the *KDM4B* Episign probe set,^39^ hierarchical clustering was performed using euclidean distance and ward.D2 on the normalized methylation values for the *FBXO22*-deficiency samples and our additional cohort of previously published ONT-seq reads from 17 unrelated Neurological Diseases (ND cohort).^45^ IGV v2.16 was used to visualize differentially methylated regions of interest from the ONT-seq reads.

### FBXO22 Epi-signature detection

In order to identify a specific FBXO22 methylation signature, we focused on regions defined by 3,643 probes from the published Episign epigenomic signature dataset^44^ for a total of 34 MNDDs. Standardized and normalized methylation values were calculated, as mentioned above. The FBXO22_F1-II:1 sample sequenced with low input DNA protocol was observed to show higher variability within the replicates, suggesting technical variability. Hence, probes with low variability within the replicates were considered. Briefly, probes with high variability within the FBXO22 sample replicates were removed such that the standard deviation of each probe of the replicates were within the 75th quantile, thus negating effects of technical variability within the replicates. Methylation difference was calculated for each probe between the MNDD and FBXO22 (i), where p is the probe, methylDiff is the methylation difference, 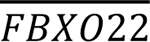 is the mean methylation value of the replicates of FBXO22 and, 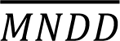 is the mean methylation value of the 34 MNDDs:

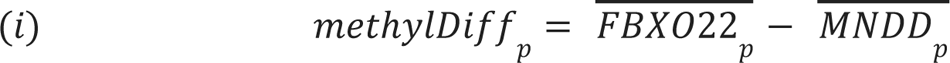

The optimal probe set for FBXO22 (40 probes) was selected by an iterative method of probe selection based on the ranked decreasing value of the methylDiff such that the cumulative explained variance for principal components 1 and 2 was at least 60% with least correlation across the MNDD.

### Haplotype and Phylogeny Analysis

ONT-seq WGS for the three samples were mapped using minimap2^76^ (v2.28-r1209) using the preset parameter (`map-ont`) to GRCh38. SNP genotype likelihoods were generated using bcftools (v1.17) using the following command on polymorphic sites found in the HGDP+APPG reference panel:
`bcftools mpileup -B -Q13 -q30 --max-BQ 30 -I -E -T chr15.reference.panel.vcf.gz -b <bam.list> -Ou | bcftools call -Aim`
`-C alleles -T chr15.reference.panel.sites.tsv.gz -Oz`

This panel is composed of the 929 HGDP samples^77^ and an additional 135 APPG (Middle Eastern) samples.^56^ The construction of the panel is described in the supplementary of Martiniano et al., 2024.^78^ Beagle4.0^79^ was subsequently used to refine genotypes based on genotype likelihoods using the HGDP+APPG reference panel. Only highly confident sites (AR2 > 0.98) were retained for analysis. Beagle5.4^80^ was then used to phase variants into haplotypes using the same reference panel. FLARE^81^ (version 0.5.1) was used to perform local ancestry inference using the HGDP samples as a reference with seven reference ancestries set as previously described^77^: Africa, Europe, Middle East, East Asia, Americas, Oceania and Central & South Asia. A phylogeny based on SNPs within the region (chr15:75794242-76113817) was generated based on fasta sequences that were produced for each haplotype using bcftools consensus (v1.17). The distance-based phylogeny (HKY) was built using seaview (v5.0.5)^83^. The three individuals were analysed along with the aforementioned 135 Middle Eastern samples. Due to the large phylogeny, only the branch with the three samples is illustrated.

### Cell Culture

The patient-derived primary cutaneous fibroblast cell line was established from a skin biopsy from F7-II:3, following standard procedures.^34,82^ All primary fibroblast lines were cultured in complete Dulbecco’s Modified Eagle Medium/High Glucose (with 4 mML-glutamine) (HyClone Cat: SH30022.01) supplemented with 10% fetal bovine serum (FBS) (Biological Industries) and 1% penicillin-streptomycin (Gibco). All cell lines were maintained in a humidified atmosphere at 5% CO_2_ and 37°C and tested negative for mycoplasma using the MycoAlert Mycoplasma Detection Kit (Lonza, catalog no. LT07-118).

### RNA Extraction and RT-qPCR

Total RNA from cell culture was extracted using the RNeasy Mini Kit (Qiagen) according to the manufacturer’s instructions. For RT-qPCR analysis, cDNA was synthesized using a ReverTra Ace qPCR kit (Toyobo). RT-qPCR amplifications were then performed in 96-well optical reaction plates with Power SYBR Green PCR Master Mix (Applied Biosystems) on the QuantStudio 3 System (Applied Biosystems). The relative expression values of each gene were determined by normalization to beta-actin expression for each sample. Prism v10 was used for statistical analysis of RT-qPCR data.

### Cell Lysate and Western Blotting

Cells were directly lysed with Laemmli-buffer (2% SDS, 10% glycerol, 5% 2-mercaptoethanol, 0.002% bromophenol blue, and 62.5 mM Tris HCl at pH 6.8). Whole lysates (20-50 g) were separated by SDS-PAGE, transferred to a PVDF (Immobilon-P; Millipore) membrane, and then subjected to immunoblotting with the appropriate antibodies using the ECL detection system.

### In silico Protein Analysis

Protein sequence conservation analysis of FBXO22 was performed with the Clustal Omega program (v1.2.4) on UniProt. AlphaFold3^33^ on the AlphaFold Server was used to model the protein structure of FBXO22 (Q8NEZ5), visualised using pyMOL (v3.0.3).

### List of antibodies used

mouse anti-ACTB (AC-15: Santa Cruz Biotechnology) rabbit anti-FBXO22 (GeneTex, GTX117774) rabbit anti-KDM4B (Cell Signaling Technology, D7E6).

### List of primers used

*ACTB*-forward primer: AGAGCTACGAGCTGCCTGAC

*ACTB*-reverse primer: AGCACTGTGTTGGCGTACAG

*FBXO22*-forward primer: CTCACTGAAGTAGGTCTTTTAG

*FBXO22*-reverse primer: CCAGCCAAGATGATATTCATATC

*KDM4B*-forward primer: TGTCTGATGAGCGTGAAAGG

*KDM4B*-reverse primer: GTTGGAGGAATCAGCCAAAA

## SUPPLEMENTAL FIGURES AND LEGENDS

**Figure S1.**
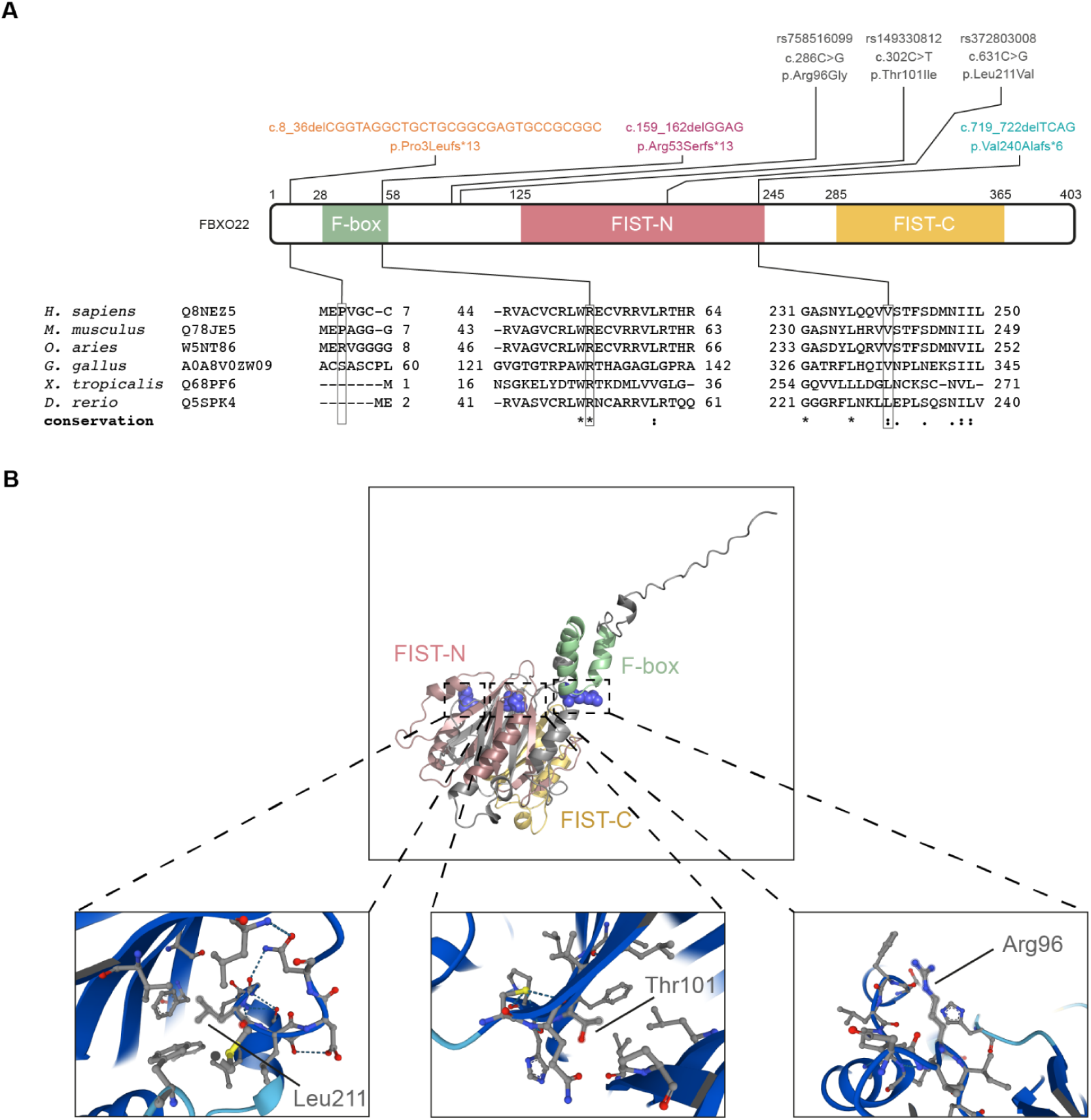
In silico analysis of *FBXO22* alleles. (A) Schematic diagram of the protein primary structure of FBXO22 with loss-of-function alleles indicated in color, and more commonly found SNPs in grey. Vertebrate sequence conservation of the amino acids implicated in the loss-of-function alleles indicated below. (B) AlphaFold 3-predicted structure of human FBXO22 (Q8NEZ5), with emphasis on the locations of the three more commonly found missense variants at Argine96 (rs758516099; c.286C>G; p.(Arg96Gly)), Threonine101 (rs149330812; c.302C>T; p.(Thr101Ile)) and Leucine211 (rs372803008; c.631C>G; p.(Leu211Val)).

**Figure S2.**
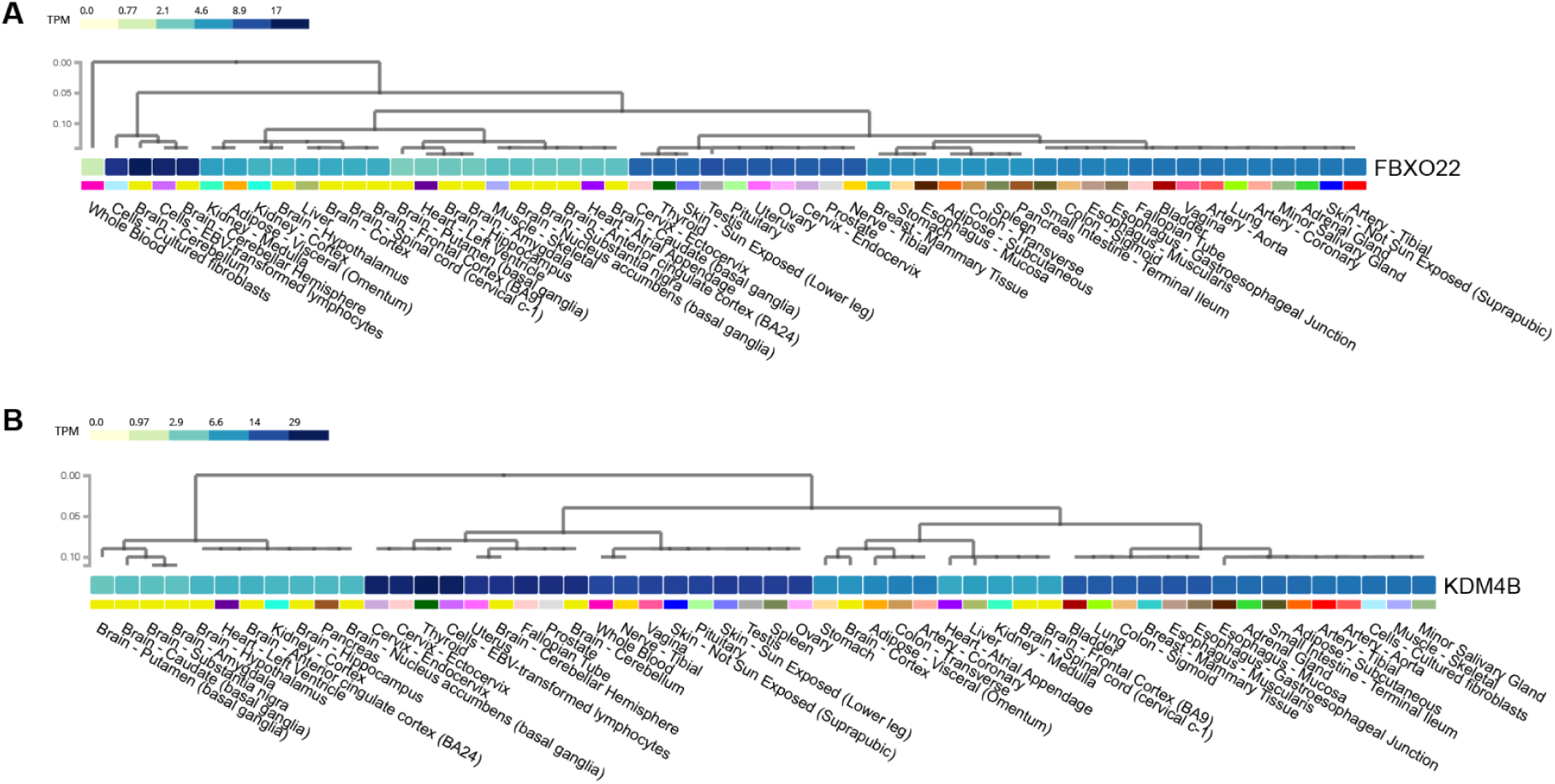
Ubiquitous expression of *FBXO22* and *KDM4B*. GTEx adult expression levels of (A) *FBXO22* and (B) *KDM4B* indicate that both genes are expressed ubiquitously over different tissue types.

**Figure S3.**
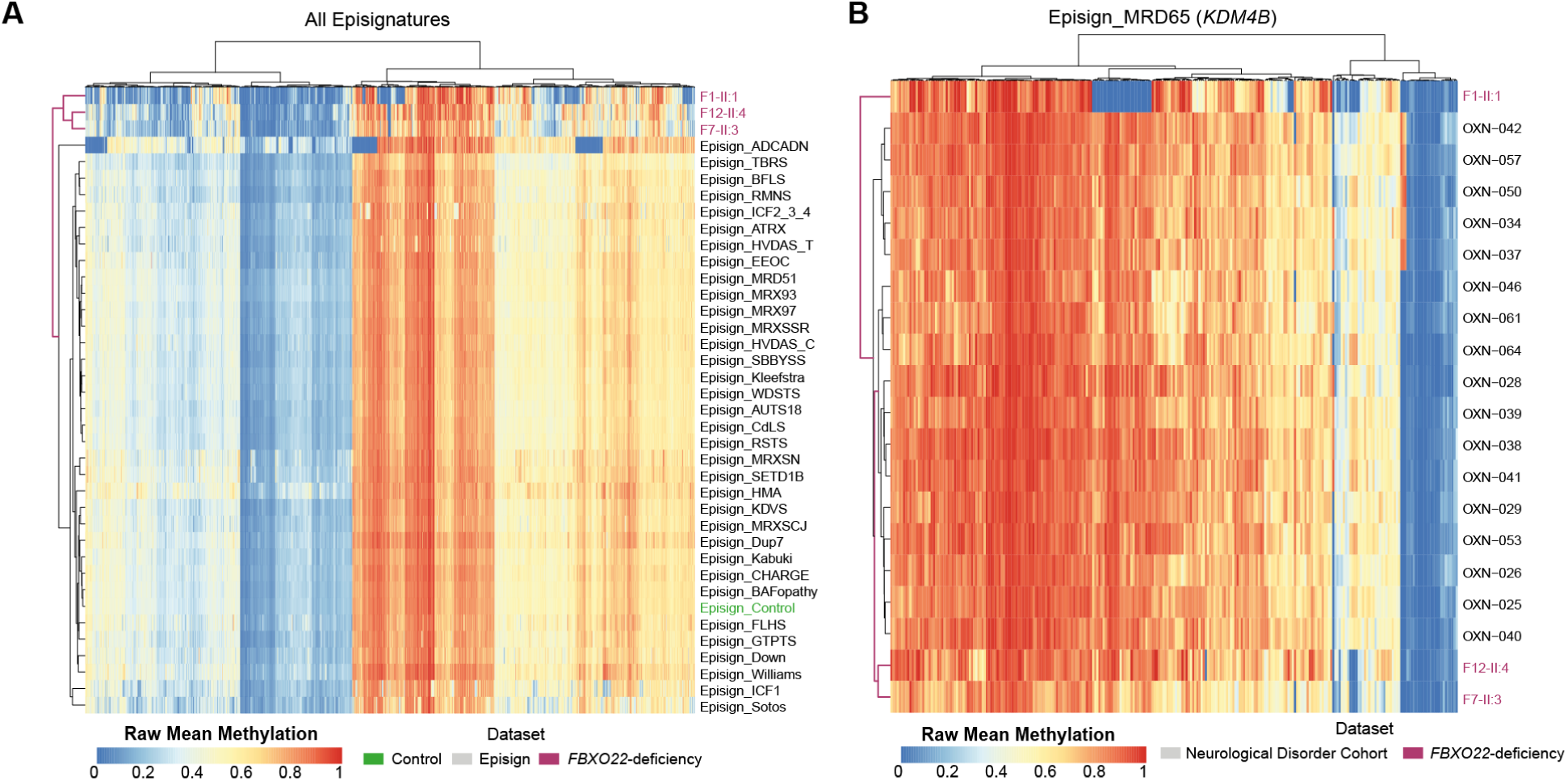
Peripheral blood DNA methylation analysis across known epi-signatures. (A) Heatmap with euclidean distance hierarchical clustering of DNA methylation values for all 3,643 regions featuring known epi-signatures for three *FBXO22*-deficiency peripheral blood samples integrated with the 34 EpiSign Mendelian neurodevelopmental disorders (MNDDs) and control dataset. (B) Heatmap with euclidean distance hierarchical clustering of methylation values across 246 regions for *KDM4B* LoF (MRD65)-defining epi-signature regions for the three *FBXO22*-deficiency samples together with an additional dataset of 17 ONT-seq samples with general neurological disorders (ND cohort).^45^

**Figure S4.**
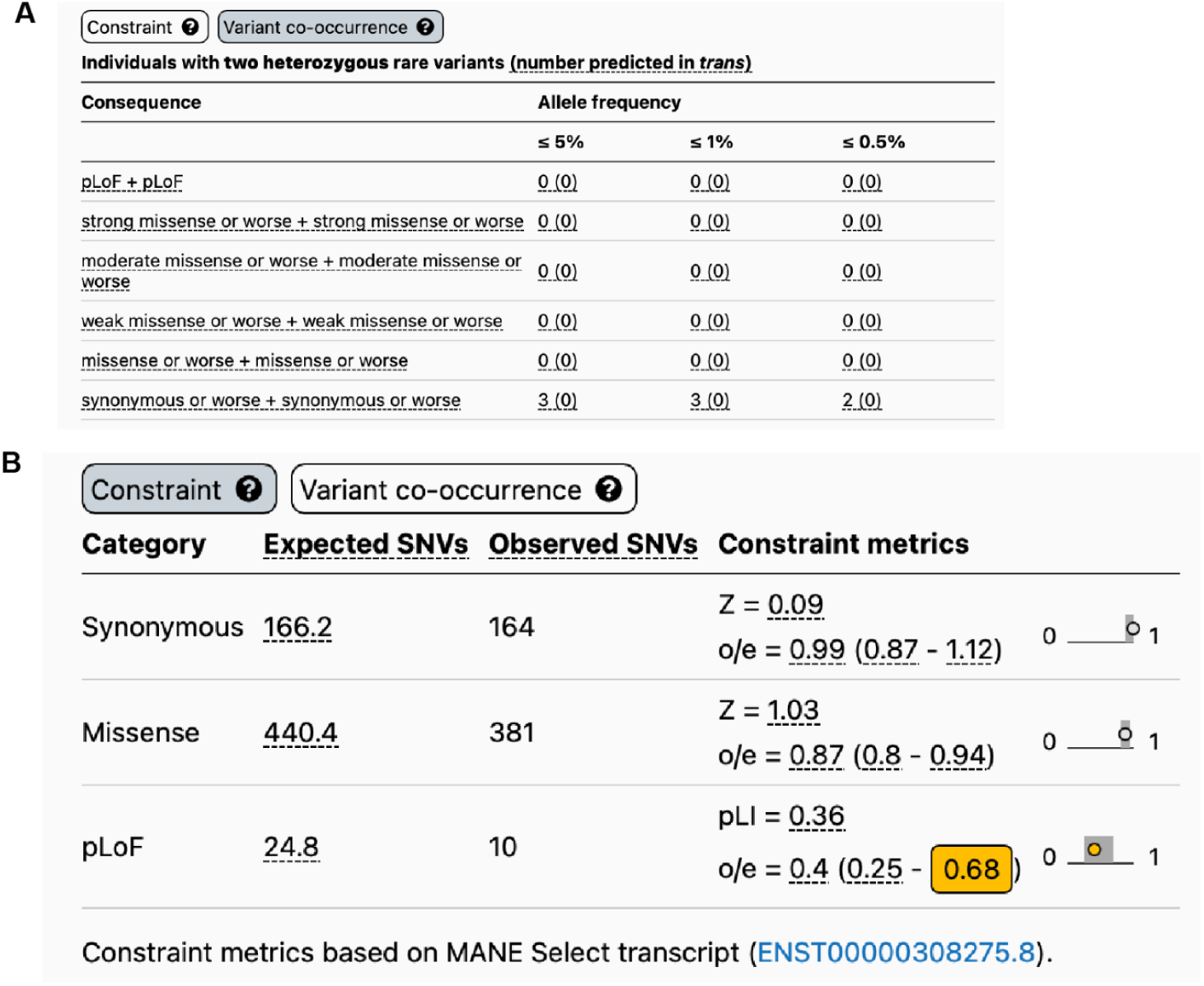
*FBXO22* intolerance to biallelic and compound heterozygous loss-of-function variants. (A) Constraint metrics on heterozygous predicted loss-of-function (pLOF) variants for *FBXO22* from gnomAD v4.1.0. (B) Heterozygous variant co-occurrence (compound heterozygous) frequencies for *FBXO22* from gnomAD v.2.1.1.

**Figure S5.**
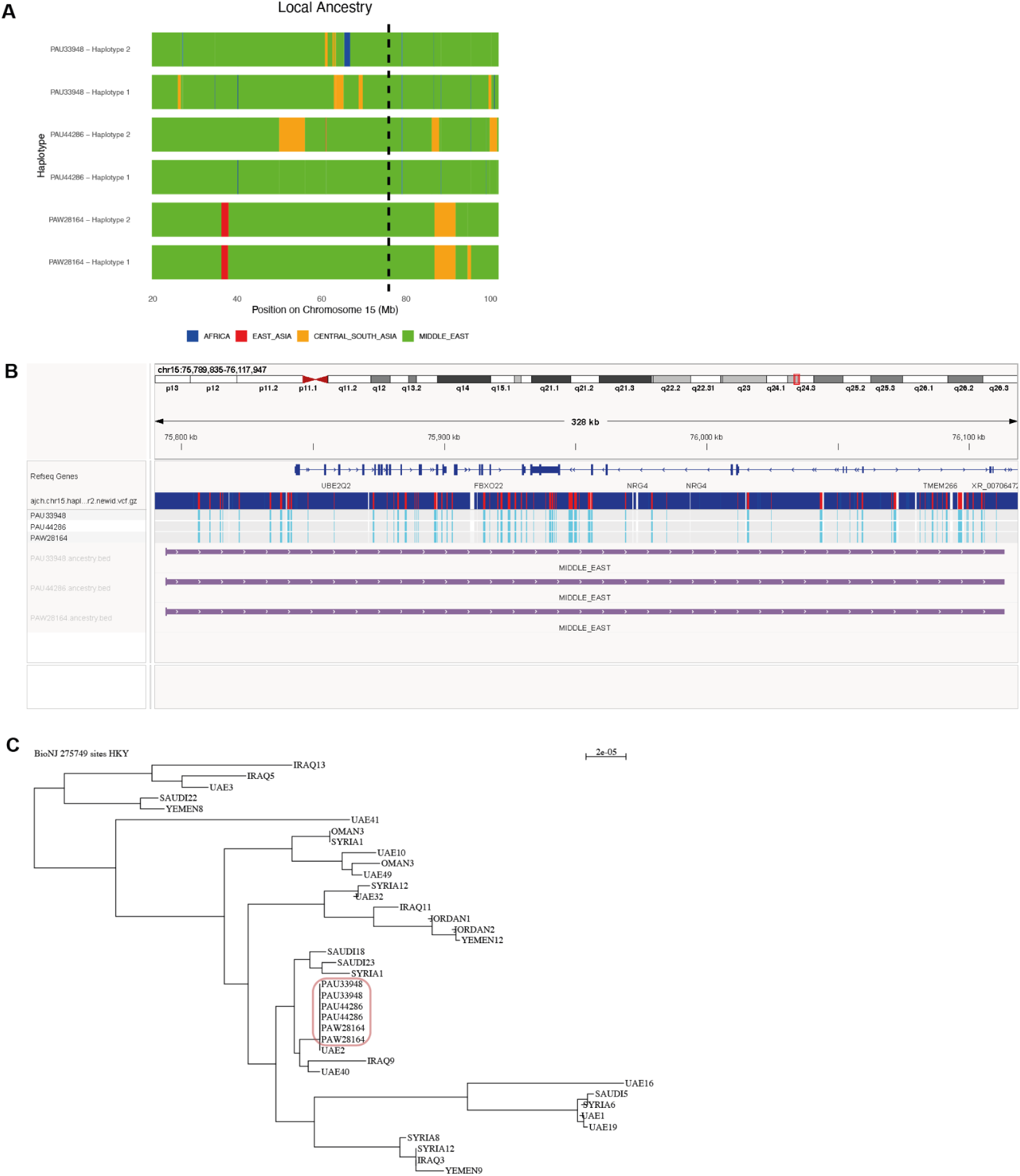
Haplotype and phylogenetic analysis of the recurrent c.159_162delGGAG; p.(Arg53Serfs*13) variant. (A) Local ancestry analysis of chromosome 15 for three ONT-seq WGS samples (see Methods for details) containing the c.159_162delGGAG; p.(Arg53Serfs*13) variant. Dashed vertical line illustrates the location of the variant. (B) hg38 genome browser view of a 328 kb region encompassing the variant identified. Light blue vertical bars indicate homozygous alternative SNPs. All three samples appear to carry an almost identical homozygous haplotype which is inferred to be of Middle Eastern origin. (C) Phylogeny of the six haplotypes in the three samples alongside 135 published Middle Eastern individuals (see Methods for details, a subset of the branch is presented here). The three samples (six haplotypes) investigated in this study cluster together (red box).

